# Steroid-Responsiveness in TBX4-Associated Pulmonary Hypertension and Interstitial Lung Disease

**DOI:** 10.64898/2026.04.19.26350630

**Authors:** Cara Morgan, Alistair Calder, Rossa Brugha, Sadia Quyam, Paul Aurora, Eimear McGovern, Andrew Bush, Shahin Moledina

## Abstract

**Background:** *TBX4* variants are a recognised cause of paediatric pulmonary hypertension (PH), often associated with interstitial lung disease (ILD). Evidence for ILD-directed therapy in this group is lacking.

**Methods:** We conducted a retrospective study of children (≤18 years) with *TBX4*-associated PH at a national centre (2001–2025). ILD was defined using ChILD-EU criteria. Patients treated with pulsed intravenous methylprednisolone were assessed for response using ChILD-EU categories. Secondary outcomes included respiratory severity score (RSS), functional class (FC), echocardiographic measures, and NT-proBNP.

**Results:** Of 21 children, 11 (52%) had ILD; 9 received corticosteroids. Median age at treatment was 0.8 years. A clear or best response occurred in 7/9 (78%). RSS improved in 6/9 (p=0.02), with all children on respiratory support showing partial or complete weaning. Functional class improved in all with FC III/IV at baseline (p=0.02). Right ventricular function improved (TAPSE z-score +1.65, p=0.04), and elevated NT-proBNP normalised. Key clinical milestones included ECMO weaning, transplant delisting, and discontinuation of prostacyclin therapy. No significant adverse effects were observed. Untreated children showed no early improvement.

**Conclusions:** Corticosteroids were associated with meaningful improvements in respiratory and PH outcomes in *TBX4*-associated PH with ILD. Prospective evaluation is warranted.

## Introduction

Pathogenic variants in *TBX4*, including point mutations and 17q23 microdeletions encompassing the gene locus, have emerged as a major monogenic contributor to paediatric pulmonary hypertension (PH).^1^ A significant proportion of children with *TBX4* associated PH (TBX4-PH) manifest features of developmental lung disease, including interstitial lung disease (ILD).^2,3^ To date, there are no data on the efficacy of ILD directed therapy in this setting. Herein, we present the first report of response to corticosteroid treatment in patients with TBX4-PH and ILD.

## Methods

We conducted a retrospective analysis of children aged ≤18 years with genetically confirmed TBX4-PH managed at a national referral centre between 2001 and 2025. Both ILD diagnosis and the decision to treat with steroids were made according to European Research Collaboration for Children’s ILD (ChILD-EU) criteria, integrating clinical features, chest imaging, multidisciplinary review and lung histology where available.^4^ Children received at least one course (range 1-6) of monthly pulsed intravenous methylprednisolone (daily 10⍰mg/kg dose for 3 days). Treatment response was graded using ChILD-EU response categories (no, probable, clear, best response).^4^ Secondary outcomes included change in respiratory severity score (RSS), World Health Organisation functional class (FC), echocardiographic indices, N-terminal pro-B-type natriuretic peptide (NT-proBNP), and six-minute walk distance where available. As PH contributes maximally to the standard ChILD-EU respiratory severity scale, an adapted RSS (range 1—5) was applied. RSS was defined as: 1, asymptomatic without hypoxaemia; 2, respiratory symptoms or signs without hypoxaemia; 3, hypoxaemia during sleep or need for nocturnal supplemental oxygen; 4, hypoxaemia at rest or requirement for continuous supplemental oxygen; and 5, need for non-invasive or invasive pressure-support ventilation. Dyspnoea, a common symptom of PH, was excluded from the score. Paired comparisons between baseline and post-treatment values were analysed using the Wilcoxon-signed rank test. The study was approved by the relevant research ethics committee (17/LO/0008); individual consent was waived.

## Results

Among 21 children with TBX4-PH, 11 (52%) met criteria for ILD (**Table 1**). Six children carried copy number variants and 5 single-nucleotide variants in *TBX4*. Nine children received pulsed intravenous methylprednisolone; therapy was declined by one family and was not offered to one child diagnosed early in the series prior to established consensus on steroid therapy in ILD. Median age at steroid initiation was 0.8 (range 0.04–14.9)⍰years. Children received between one and six pulses. No significant steroid-related adverse effects were observed. All children receiving steroids were established on background pulmonary vasodilator therapy at baseline, most commonly sildenafil (n=8) with or without bosentan (n=6); four were receiving prostanoid therapy (inhaled =2, intravenous = 2). One child was treated with inhaled nitric oxide as a single agent. No new pulmonary vasodilators were initiated during steroid treatment period. Concomitant non-steroid ILD-directed therapies included azithromycin (n=8) and hydroxychloroquine (n=4), and 2 children were treated with inhaled corticosteroids. Overall, 7/9 children (78%) achieved a clear or best response, one had a probable response, and one had no response. Respiratory severity score improved in 6 of 9 (67%) children (p=0.02). All children requiring continuous respiratory support at baseline (n=8) demonstrated partial or complete weaning during or following treatment, including children requiring invasive ventilation (n=2) or extracorporeal membrane oxygenation (n=1).

**Table 1:** Response to steroid treatment in children with TBX4 associated pulmonary hypertension and interstitial lung disease. **Key**: BiPAP = biphasic positive airway pressure support; CNV = copy number variant inclusive of TBX4; ECMO = extra-corporeal membrane oxygenation; FC = functional class; HF O_2_; high flow oxygen therapy; IV = intravenous; LF O_2_ = low flow oxygen therapy; NT-proBNP = N terminal pro B-type natriuretic peptide; O_2_ = oxygen therapy; PH = pulmonary hypertension; PPHN = persistent pulmonary hypertension of the newborn; SNV = single nucleotide variant in TBX4; TAPSE = tricuspid annular plane systolic excursion; TR Vmax = peak tricuspid regurgitant jet velocity; 6MWD = six-minute walk distance. *Established on inhaled iloprost added between 5^th^ and 6^th^ steroid pulse.

| Age at Baseline assessment<br>*Diagnosis* | Variant | Pulmonary vasodilator | Additional therapy | No. pulses | Respiratory support |  | Respiratory response | FC |  | Changes in NT-pro-BNP, 6MWD, and echocardiographic assessment of PH at early follow-up | Change to PH therapy |
| --- | --- | --- | --- | --- | --- | --- | --- | --- | --- | --- | --- |
|  |  |  |  |  | Baseline | post |  | Baseline | post |  |  |
| Children receiving steroid therapy |  |  |  |  |  |  |  |  |  |  |  |
| 0-6 months<br>*PPHN* | CNV | Inhaled nitric oxide | Nil | 1 | VV ECMO | 24hr LF O2 | Best response | 4 | 2 | Echocardiographic changes: TAPSE z-score -3.7 to -3.5; TR Vmax 3.7 m/s to 3.1 m/s | Weaned from VV ECMO/iNO |
| 0-6 months<br>*PPHN* | CNV | Sildenafil, bosentan, inhaled iloprost, amlodipine | Azithromycin | 6 | HF + O2 | 24hr LF O2 | Response | 4 | 2 | NT-proBNP: 6,990 to 314 pg/ml. Echocardiographic changes: TAPSE z-score -2.6 to -1.2; TR Vmax 3.9 m/s to insufficient. | None |
| 0-5 months<br>*PPHN* | SNV | Sildenafil | Azithromycin | 1 | HF + O2 | 24hr LF O2 | Response | 2 | 2 | Echocardiographic changes: TAPSE z-score -4.3 to -3.8; TR Vmax 4.1 to 3.9 m/s. | None |
| 0-5 mnths<br>*PPHN* | CNV | Sildenafil, bosentan | Azithromycin, hydroxychloroquine | 6 | Ventilated | BiPAP + O2 | Best response | 4 | 3 | NTpro-BNP: 11,331 to 205 pg/ml. Echocardiographic changes: TAPSE z-score +1.1 to +1.9; TR Vmax 4.0 to 4.5 m/s; | None |
| 6-11 months<br>6-11 months | SNV | Sildenafil, bosentan, IV epoprostenol | Azithromycin, hydroxychloroquine | 6 | Ventilated | 24hr LF O2 | Best response | 4 | 3 | NT-proBNP: 231 to 203 pg/ml. Echocardiographic changes: TAPSE z-score -6.1 to -3.0; TR insufficient. | Weaned from IV epoprostenol during steroid course. |
| 1-5 years<br>1-5 years | SNV | Sildenafil, nifedapine | Azithromycin, Fluticasone propionate | 3 | None | None | Probable response | 2 | 1 | NT-proBNP: 332 to 260 pg/ml. Echocardiographic changes: TAPSE z-score -3.7 to 2.4; TR Vmax 4.2 to 4.3 m/s. | None |
| 1-5 years<br>1-5 years | SNV | Sildenafil, bosentan, | Azithromycin, hydroxychloroquine | 6 | HF + O2 | 24hr LF O2 | Best response | 4 | 2 | NT-proBNP: 35,000 to 276 pg/ml. Echocardiographic changes: | Weaned from inhaled iloprost at |
|  |  | inhaled<br>iloprost* |  |  |  |  |  |  |  | TAPSE z-score -6.9 to -0.4;<br>TR Vmax 4.3 to 2.4 m/s. | 4.2 years of age.<br>Lung transplant de-<br>listing. |
| **11-15 years**<br>*1-5 years* | SNV | Sildenafil,<br>bosentan, IV<br>epoprostenol | Fluticasone<br>propionate | 6 | Nocturnal<br>LF O2 | Nocturnal<br>LF O2 | None | 3 | 2 | 6MWD increased from 440 to 560m<br>Echocardiographic changes:<br>TAPSE z-score -0.4 to +3.1;<br>TR Vmax 3.2 to 3.5 m/s. | Weaned from IV<br>epoprostenol at 12.2<br>years of age. |
| **11-15 years**<br>*0-5 months* | CNV | Sildenafil,<br>bosentan, IV<br>epoprostenol | Azithromycin<br>Hydroxychloroquine | 3 | 24hr LF<br>O2 | Nocturnal<br>LF O2 | Response | 3 | 3 | Echocardiographic changes:<br>TAPSE z-score -2.5 to -5.2;<br>TR Vmax 5.5 to 5.0 m/s. | None |
| **Children not receiving steroid therapy** |  |  |  |  | **Baseline** | **F/U**<br>**< 1 year** |  | **Baseline** | **F/U**<br>**< 1 year** |  |  |
| **16-20 years**<br>*2.7 years* | CNV | Sildenafil | Azithromycin | None | None | None | N/A | 2 | 2 | Echocardiographic changes:<br>TAPSE z-score -2.2 to -2.7;<br>TR Vmax 4.2 o 4.2 m/s. | None |
| **1-5 years**<br>*6-11 months* | CNV | Sildenafil,<br>bosentan, IV<br>epoprostenol | None | None | 24 hr LF<br>O2 | 24hr LF<br>O2 | N/A | 2 | 2 | Echocardiographic changes:<br>TAPSE z-score n/a to -3.8;<br>TR Vmax incomplete to 5.5 m/s. | None |

Functional class improved significantly (p=0.02); all children in FC III/IV at baseline (n=7) improved, while those in FC II (n=2) remained stable. There was a significant improvement in tricuspid annulus plane systolic excursion (z-score change: +1.65, p = 0.04). NT-proBNP concentrations were elevated at baseline in three children and normal in two; elevated values normalised during treatment, and normal values were maintained (**Figure 1**). In the one child able to perform serial six-minute walk testing, distance increased from 440 m to 560 m following treatment.

**Figure 1:**
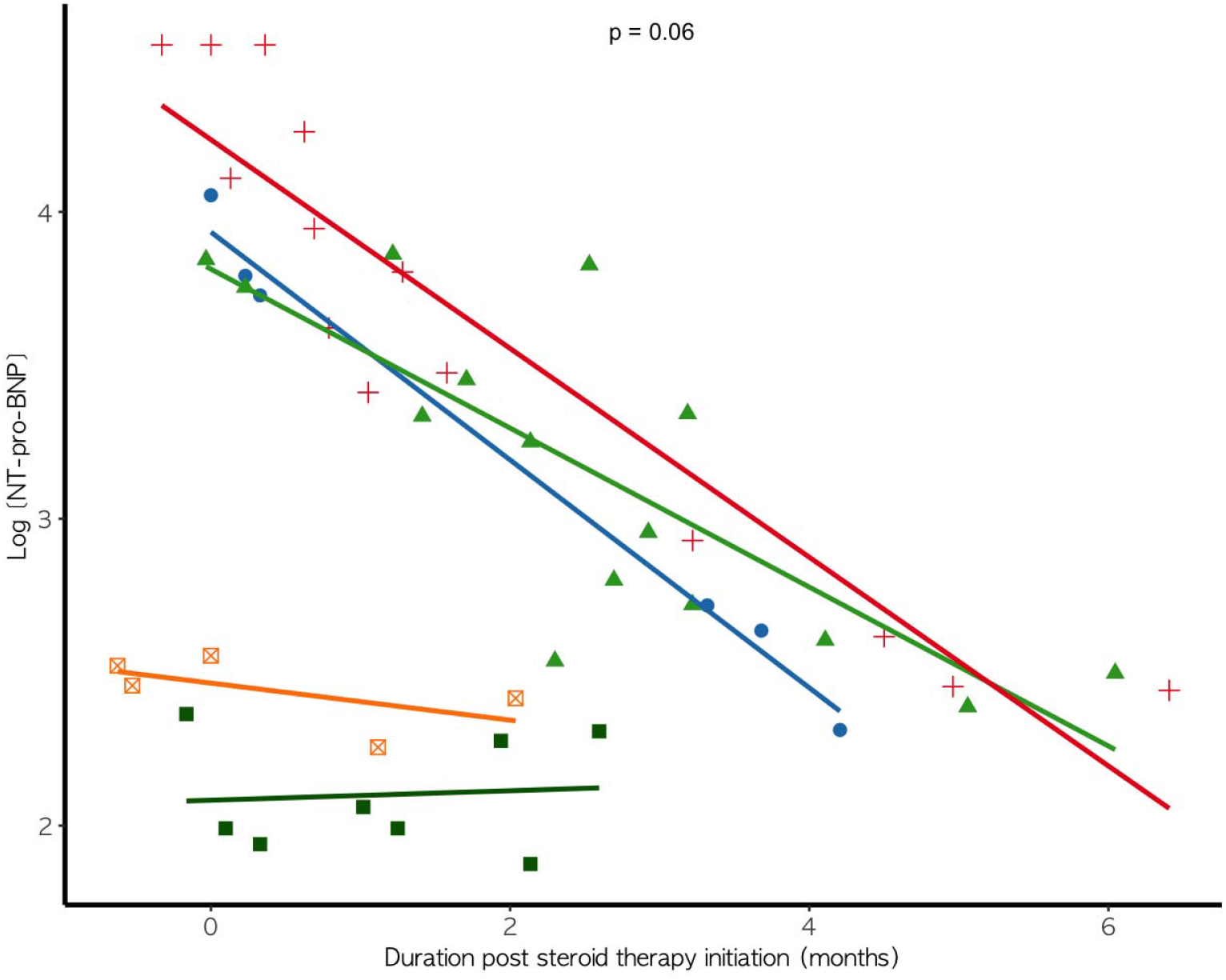
Change in NT-proBNP levels in 5 children with TBX4 associated pulmonary hypertension and interstitial lung disease who were treated with a course of pulsed intravenous methylprednisolone (6-month course = 3, 3-month course = 2).

Steroid treatment coincided with key milestones of clinical improvement in 5 children: one with severe persistent pulmonary hypertension of the newborn was weaned from extra-corporeal membrane oxygenation and inhaled nitric oxide shortly after a single steroid pulse; one child listed for lung transplantation was delisted following marked improvement in both lung disease and PH; prostacyclin therapy was discontinued in 3 children at a median 17 (range 4—25) months after steroid initiation. Over a median followup of 3.3 (range 0.7–10.4) years, no deaths occurred and one child underwent bilateral lung transplantation. Among the remaining 8 children, the median respiratory severity score was 1 (range 1–4), and all were in FC I/II at last follow-up.

In contrast to the steroid-treated group, the two children who did not receive steroids remained in FC II with no echocardiographic change at early follow-up (<1 year); respiratory support was unchanged in the one child who required it.

## Discussion

Our observations suggest that corticosteroid therapy may yield clinically meaningful improvements in both respiratory status, functional class, and markers of PH severity in children with TBX4-PH and ILD. The uncontrolled and retrospective nature of this study precludes causal inference, and treatment decisions were made clinically. Nonetheless, the consistency and magnitude of observed improvements across respiratory and pulmonary vascular disease warrants further evaluation in this subgroup of patients. Prospective study through a multicentre registry or small-n study is needed to assess safety and explore potential efficacy of intravenous methylprednisolone, which may alter management paradigms for children with TBX4-assosciated lung disease.

## Data Availability

All data produced in the present work are contained in the manuscript

## Acknowledgements

We sincerely thank all clinicians associated with the UK Pulmonary Hypertension Service for Children, who share the care for these complex patients. Without their cooperation and collaborative work, this study would not have been possible. The authors thank the Dinosaur Trust and Great Ormond Street Children’s Charity for supporting research capacity within the Paediatric Pulmonary Hypertension Unit. All research at Great Ormond Street Hospital NHS Foundation Trust and University College London Great Ormond Street Institute of Child Health is made possible by the NIHR Great Ormond Street Hospital Biomedical Research Centre. The views expressed are those of the author(s) and not necessarily those of the NHS, the NIHR or the Department of Health. This research received no specific grant from any funding agency in the public, commercial, or not-for-profit sectors.

## Conflicts of interest disclosure

Shahin Moledina has acted as a consultant for Janssen-Cilag ltd and GSK. Cara Morgan, Alistair Calder, Rossa Brugha, Sadia Quyam, Paul Aurora, Eimear McGovern, and Andrew Bush have nothing to declare.

## Notes

**“At a Glance”:** Children with TBX4-associated pulmonary hypertension and interstitial lung disease showed meaningful improvements in respiratory status and pulmonary vascular disease following pulsed intravenous corticosteroids, supporting further prospective evaluation.

### Funding Statement

This study did not receive any funding

### Author Declarations

This study was approved by London - South East Research Ethics Committee (17/LO/0008) allowing individual consent to be waived.

## References

1. Welch CL, Austin ED, Chung WK. Genes that drive the pathobiology of pediatric pulmonary arterial hypertension. Pediatr Pulmonol. 2021;56(3):614–620. doi:10.1002/ppul.24637

2. Galambos C, Mullen MP, Shieh JT, et al. Phenotype characterisation of TBX4 mutation and deletion carriers with neonatal and paediatric pulmonary hypertension. European Respiratory Journal. 2019;54(2):1801965. doi:10.1183/13993003.01965-2018

3. Karolak JA, Vincent M, Deutsch G, et al. Complex Compound Inheritance of Lethal Lung Developmental Disorders Due to Disruption of the TBX-FGF Pathway. The American Journal of Human Genetics. 2019;104(2):213–228. doi:10.1016/j.ajhg.2018.12.010

4. Bush A, Cunningham S, de Blic J, et al. European protocols for the diagnosis and initial treatment of interstitial lung disease in children. Thorax. 2015;70(11):1078–1084. doi:10.1136/thoraxjnl-2015-207349

